# Immune and pathophysiologic profiling of antenatal COVID-19 in the GIFT cohort: A Singaporean case-control study

**DOI:** 10.1101/2022.04.19.22273864

**Authors:** Yue Gu, Jia Ming Low, Jolene S.Y. Tan, Melissa Shu Feng Ng, Lisa F.P. Ng, Bhuvaneshwari D/O Shunmuganathan, Rashi Gupta, Paul A. MacAry, Zubair Amin, Le Ye Lee, Derrick W.Q. Lian, Lynette Pei-Chi Shek, Youjia Zhong, Liang Wei Wang

**Affiliations:** Antibody Engineering Programme, Life Sciences Institute, National University of Singapore, Singapore; Department of Microbiology and Immunology, Yong Loo Lin School of Medicine, National University of Singapore, Singapore; Department of Neonatology, Khoo Teck Puat-National University Children’s Medical Institute, National University Health System, Singapore; Department of Paediatrics, Yong Loo Lin School of Medicine, National University of Singapore, Singapore; Duke-NUS Medical School, Singapore; Singapore Immunology Network, Agency for Science, Technology and Research, Singapore; Infectious Diseases Labs, Agency for Science, Technology and Research, Singapore; Department of Pathology, National University Hospital, Singapore; Khoo Teck Puat-National University Children’s Medical Institute, National University Health System, Singapore

## Abstract

**Background:** COVID-19 has been a major public health threat for the past two years, with disproportionate effects on the elderly, immunocompromised, and pregnant women. While much has been done in delineating immune dysfunctions and pathogenesis in the former two groups, less is known about the disease’s progression in expectant women and children born to them. To address this knowledge gap, we profiled the immune responses in maternal and child sera as well as breast milk in terms of antibody and cytokine expression and performed histopathological studies on placentae obtained from mothers convalescent from antenatal COVID-19.

**Methods and findings:** A total of 17 mother-child dyads (8 cases of antenatal COVID-19 and 9 healthy unrelated controls; 34 individuals in total) were recruited to the Gestational Immunity For Transfer (GIFT) study. Maternal and infant sera, and breast milk samples were collected over the first year of life. All samples were analyzed for IgG and IgA against whole SARS-CoV-2 spike protein, the spike receptor-binding domain (RBD), and previously reported immunodominant epitopes, with conventional ELISA approaches. Cytokine levels were quantified in maternal sera using multiplex microbead-based Luminex arrays. The placentae were examined microscopically. We found high levels of virus-specific IgG in convalescent mothers and similarly elevated titers in newborn children. Virus-specific IgG in infant circulation waned within 3-6 months of life. Virus-specific IgA levels were variable among convalescent individuals’ sera and breast milk. Convalescent mothers also showed a blood cytokine signature indicative of a persistent pro-inflammatory state. Four placentae presented signs of acute inflammation marked by neutrophil infiltration even though >50 days had elapsed between virus clearance and delivery. Administration of a single dose of BNT162b2 mRNA vaccine to mothers convalescent from antenatal COVID-19 increased virus-specific IgG and IgA titers in breast milk.

**Conclusions:** Antenatal SARS-CoV-2 infection led to high plasma titres of virus-specific antibodies in infants postnatally. However, this was not reflected in milk; milk-borne antibody levels varied widely. Additionally, placentae from COVID-19 positive mothers exhibited signs of acute inflammation with neutrophilic involvement, particularly in the subchorionic region. Virus neutralisation by plasma was not uniformly achieved, and the presence of antibodies targeting known immunodominant epitopes did not assure neutralisation. Antibody transfer ratios and the decay of transplacentally transferred virus-specific antibodies in neonatal circulation resembled that for other pathogens. Convalescent mothers showed signs of chronic inflammation marked by persistently elevated IL17RA levels in their blood. A single dose of the Pfizer BNT162b2 mRNA vaccine provided significant boosts to milk-borne virus-specific antibodies, highlighting the importance of receiving the vaccine even after natural infection with the added benefit of enhanced passive immunity. The study is registered at clinicaltrials.gov under the identifier NCT04802278.

## Introduction

Severe acute respiratory syndrome coronavirus 2 (SARS-CoV-2), the causative agent of coronavirus disease 2019 (COVID-19), has infected over 497 million worldwide (WHO website, as of 13^th^ April 2022) and led to a pandemic. In the midst of this unprecedented crisis, studies on vulnerable groups such as pregnant women and newborns are limited compared to the general adult population. Although the manifestation of COVID-19 is less severe in the paediatric population compared to adults, infants are particularly susceptible to developing severe illness [1]. Additionally, concerns over mother-to-child transmission of the virus have led to variable recommendations on postnatal care [2]. Human breast milk (BM) is the main source of nutrients and bioactive factors that protects infants against general infections [3]. Maternal antibodies which are present in abundance in BM are able to confer protection against specific infections through their antigen specificity. These maternal antibodies comprise of approximately 90% immunoglobulin A (IgA), 8% IgM and 2% IgG [4]. SARS-CoV-2 antibodies from convalescent COVID-19 plasma have been extensively studied as a therapeutic option against COVID-19 infection [5,6]. Several studies have also reported the presence of SARS-CoV-2 antibodies in the BM of convalescent COVID-19 mothers [7-9]. However, the durability of those antibodies in the BM and the mechanism of protection remains incompletely known. Hence, we sought to evaluate the durability and neutralization capacity of SARS-CoV-2 specific IgG and IgA in the BM of convalescent COVID-19 mothers.

Here, we report that natural infection of pregnant women by SARS-CoV-2 induced antibody production and secretion into maternal blood and milk. However, plasma antibody titres were not closely reflected by that in milk. Placentae from COVID-19 positive mothers exhibited signs of acute inflammation with neutrophilic involvement, despite virus clearance in the nasopharynx. Virus neutralisation by plasma was not uniformly achieved, despite high levels of antibodies targeting the spike protein and its receptor-binding domain; the presence of antibodies targeting known immunodominant epitopes did not assure neutralisation. Antibody transfer ratios and decay kinetics of virus-specific antibodies in neonatal circulation largely resembled that described for other pathogens. Targeted analysis of blood cytokines revealed significant elevations in IL17RA levels in convalescent mothers’ blood at 16 weeks (range 13-48) from the initial COVID-19 infection, indicating chronic inflammation. Importantly, we also found that a single regular dose of the Pfizer BNT162b2 mRNA vaccine given postnatally boosted milk-borne virus-specific IgG and IgA in mothers convalescent from antenatal COVID-19.

## Materials and methods

### Ethics statement

This study was approved by the National Healthcare Group Institutional Review Board (Gestational Immunity For Transfer GIFT: DSRB Reference Number: 2020/00483). Written informed consent was obtained from all subjects (and where applicable, parents), and the study was conducted in accordance with the Helsinki Declaration. The study protocol was registered at clinicaltrials.gov (NCT04802278).

### Collection of human breast milk

Breast milk from study participants were collected at 1 month and 3 months postpartum. Milk samples were aliquoted and frozen at -20 degrees Celsius until use. Recruited mothers who were convalescent from antenatal COVID-19 (n=8 provided blood samples; n=6 provided BM samples) were confirmed to be positive for COVID-19 infection with real-time reverse-transcriptase polymerase-chain-reaction (RT-PCR) assay on nasopharyngeal swabs during pregnancy. Recovery was defined by the resolution of clinical symptoms and two negative SARS-CoV-2 RT-PCR swabs 24 hours apart. Infants born to antenatal COVID-19 convalescent mothers at gestational age between 35 to 40 weeks gestation were included. Mother-child control dyads (n=10) were also recruited. These control mothers had no clinical symptoms of COVID-19 infection and were confirmed to have a negative SARS-CoV-2 IgG at the time of recruitment.

### Blood processing for plasma

Venous blood samples from the infant were collected at 0-3 days, 1 month, and 3 months of life, and from the mothers at 1 month postpartum, in BD Vacutainer cell preparation tubes (BD, #362753). The plasma fraction was harvested after centrifugation at 1500rpm, 5 minutes at 4°C and stored at -80°C. Sample viral inactivation was performed by treatment with 1% Triton X-100 (Thermo Fisher Scientific, #28314) and 0.3% (w/w) tri-(*n*-butyl) phosphate (TNBP) at room temperature for 2 hours.

### Synthesis of the SARS-CoV-2 receptor binding domain (RBD) and spike protein

SARS-CoV-2 spike and RBD were generously provided by the Antibody Engineering Programme, Life Sciences Institute, NUS as described previously [10].

### Spike and RBD-protein based ELISAs

IgA and IgG against SARS-CoV-2 antigens including the whole spike and RBD protein were titrated using quantitative ELISA. 96-well flat-bottom maxi-binding immunoplate (SPL Life Sciences, #32296) were coated with 100 ng of SARS-CoV-2 whole spike protein or 200ng of RBD protein at 4 °C overnight. After three washes in Phosphate Buffer Saline (PBS), 350 µL of blocking buffer [4% skim milk in PBS with 0.05% Tween 20 (PBST)] was added to each well. After incubation for 1.5 hours, the plate was washed three times with PBST. 100 µL of 10-times diluted human milk samples, or 100-times diluted human plasma samples were added to each well for 1-hour incubation. Plate was then washed three times with PBST followed by 1-hour incubation in the dark with 100 µL of 5000-times diluted goat anti-human IgG-HRP (Invitrogen, #31413), or 5000-times diluted F(ab’)2 anti-human IgA-HRP (Invitrogen, #A24458). Plate was washed three times in PBST and incubated for 3 minutes with 1-Step Ultra TMB-ELISA (Thermo Scientific, #34029), 100 µL per well. Reaction was stopped with 100 µL of 1 M H2SO4 and optical density at 450nm (OD450) was measured using a microplate reader (Tecan Sunrise). OD450 was calculated by subtracting the background signal from sample binding to the blocking buffer. Experiments were performed at least three times.

### Pseudovirus neutralization test (PVNT)

The PVNT assay was conducted as described previously [11]. Briefly, the ACE2 stably expressed CHO cells were cultured at 5 × 10^4^/mL cells in complete medium for 24 hours. 80-times diluted plasma were incubated with 50,000 lentiviral particles (representative of the SARS-CoV-2 Wuhan-Hu-1 strain) in a total volume of 50 µL at 37 °C for 1 hour. This mixture was added to the CHO-ACE2 monolayer cells and left incubated for 1 hour to allow pseudotyped viral infection. Subsequently, complete medium was added at 150 µL/well for further incubation of 48 hours. After two washes with PBS, 100 µL of ONE-gloTM EX luciferase assay reagent (Promega, #E8130) was added to each well and the luminescence values were recorded. The percentage neutralization was calculated as follows:

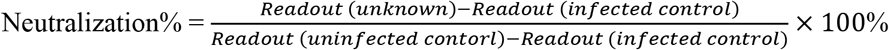

### Breastmilk/plasma inactivation and peptide based ELISA

Both plasma and BM samples were inactivated with Triton™ X-100 (ThermoFisher Scientific, #28314) to a final concentration of 1% for 2 hours at room temperature (RT). A focused epitope screen was performed according to a previously described peptide-based ELISA [12]. S14P5, S20P2, S21P2 and N4P5 are 18-mer peptides derived from SARS-CoV-2 spike protein with immunodominant activities [12,13]. For BM ELISA, Streptavidin coated plates (Life Technologies Pierce, #15126) were coated overnight at 4 degrees with 10 µg/ml peptides diluted in 0.01% polyvinyl alcohol (PVA). Plates were blocked with 0.01% PVA at RT prior to the addition of BM at 1:10 dilution. Goat HRP-conjugated anti-human IgA (Abcam, #ab97215) diluted at 1: 2,000 in the blocking buffer was used for the detection of peptide specific antibodies. TMB substrate (eBioscience, #00-4201-56) was added to the plate for development and the reaction was quenched with 2M sulfuric acid. Absorbance measurements were read at two wavelengths (450nm and 570nm) using the Infinite M200 plate reader (Tecan, firmware V_2.02_11/06). Plates were incubated at RT for one hour on a rotating shaker for all steps unless otherwise stated and washed thrice with 0.1% PBST in between steps.

For plasma ELISA, Maxisorp plates (Thermofisher Scientific, #442404) were coated overnight at 4 degrees with 1 µg/ml of Neutravidin (ThermoFisher Scientific, #31050) diluted in PBS. Plates were blocked with 0.01% PVA followed by the addition of 1:100 inactivated plasma samples. The subsequent steps followed the same sequence as mentioned above.

### Multiplex microbead-based immunoassay

Quantification of cytokine levels in the plasma samples of convalescent and healthy mothers was performed by multiplex microbead-based immunoassays. Plasma samples were treated with 1% Triton™ X-100 solvent detergent for virus inactivation [14]. Concentrations of immune mediators were determined using the Luminex™ assay (HCYTOMAG-60K-41plex) (Millipore Merck). Plasma from participants (n=7 from each group) and standards were incubated with fluorescent-coded magnetic beads pre-coated with capture antibodies in a 96-well plate. Biotinylated detection antibodies were incubated with the cytokine-bound beads for an hour. Streptavidin-PE was then added for another 30 mins before the acquisition of data using xPONENET^®^ 4.0 (Luminex Corporation, USA) software. Data was analysed using the Bio-Plex Manager™6.1.1. Standard curves were generated with a 5-PL (5-parameter logistic) algorithm, reporting values of median fluorescence intensity (MFI) and concentration data.

Real time reverse transcriptase polymerase chain reaction (RT-PCR) for COVID-19 PCR swabs were obtained from amniotic fluid, umbilical cord, placental, umbilical cord blood, maternal blood and high vaginal swabs and breast milk [15,16].

The placenta specimen was fixed in formalin overnight before representative sections were obtained from the umbilical cord, placental membranes and placental disc. The tissue was subsequently processed and stained with hematoxylin and eosin for microscopic examination.

### Statistical Analysis

The OD of samples obtained by subtracting the background signals and normalizing the samples against a negative control to account for interpolate variations. Data analyses were performed using GraphPad Prism (GraphPad Software, version 7.0.0). Unless otherwise stated, statistical significance is defined as p-values being less than 0.05.

## Results

### Clinical demographics of the GIFT cohort

Eight women with a diagnosis of laboratory confirmed COVID-19 during pregnancy participated in the study (S1 Table). Nine women with no antenatal COVID-19 were recruited as controls. On average, COVID-19 convalescent women were 30.3 ± 2.4 years old. Additional characteristics of the study participants and their infants are presented in Table 1. Of the 8 women, 6 had mild COVID-19, with the most common being dry cough, sore throat and malaise, with one having moderate COVID-19 and the remaining person being asymptomatic. None of them progressed towards hypoxia or exhibited signs of lower respiratory tract infection (LRTI) evidenced by chest X-ray abnormalities. Consequently, none of the subjects required intensive care unit admission, had oxygen requirements, or were intubated. Individual clinical demographics of the convalescent women are listed in S2 Table. Extensive testing of placental and umbilical cord samples as well as infant throat swabs all produced negative PCR results for COVID-19 (detailed reporting of results in S3 Table).

**Table 1.** Demographic and clinical characteristics of the cohort.

| **Maternal Characteristics** | **Cases (n=8)** | **Controls (n=9)** |
| --- | --- | --- |
| Age (average; years) (standard deviation, SD) | 30.3 ± 2.4 | 32.1 ± 2.5 |
| **Race/ethnicity:** |  |  |
| Chinese | 1 | 6 |
| Malay | 3 | 2 |
| Indian | 2 | 1 |
| Caucasian | 2 | 0 |
| BMI (SD) | 27 ± 2.2 | 25.7 ± 1.5 |
| No past medical history | 7 | 8 |
| Other co-morbidities if any |  |  |
| - Hepatitis C carrier | 1 | 2 |
| - Gestational diabetes mellitus (GDM) | 0 | 1 |
| - Smoking | 1 | 1 |
| - Pre-eclampsia | 0 | 0 |
| Antenatal fetal anomaly scan | No abnormalities detected | No abnormalities detected |
| Occurrence of mastitis during the study period | 0 | 0 |
| **Timing of COVID-19 infection:** |  |  |
| First trimester | 2 | - |
| Second trimester | 2 | - |
| Third trimester | 4 | - |
| **Severity** |  |  |
| Symptomatic | 6 mild, 1 moderate | - |
| Asymptomatic | 1 | - |

| **Infant Characteristics** | **Cases (n=8)** | **Controls (n=9)** |
| --- | --- | --- |
| Female | 6 | 6 |
| Gestational age when born (weeks) (SD) | 39 ± 2 | 39 ± 1 |
| **Growth at birth:** |  |  |
| Birth weight (kg) (SD) | 3.517 ± 0.261 | 3.108 ± 0.376 |
| Length (cm) (SD) | 50.5 ± 1.2 | 50.6 ± 1.3 |
| Occipital frontal circumference (cm) (SD) | 34.5 ± 0.7 | 33.3 ± 0.7 |
| **Breastfeeding status:** |  |  |
| Exclusive | 6 | 9 |
| Mixed feeding | 2 | 0 |

The eight infants were all born full term with mean gestational age (SD) of 39 ± 2 weeks and had a mean birth weight (SD) of 3.517 ± 0.261 kg. One infant required neonatal intensive care unit stay for 2 days for transient tachypnea of the newborn and was given supplemental oxygen therapy. All infants were breastfed up to 3 months, two of whom were supplemented with formula milk during the same period.

It remains controversial whether SARS-CoV-2 can be vertically transmitted and if so, what damage it might cause to the developing child and the maternal-foetal interface i.e., the placenta, especially in continued pregnancy post convalescence from antenatal COVID-19 [17-19]. To address that question, histological examination of placental samples was done to determine if there were notable pathologies or abnormalities. While all 8 samples did not show the presence of SARS-CoV-2, three placentae (CS02, CS03, CS07) showed signs of acute inflammation (Fig 1). Neutrophils were seen in the subchorionic fibrin, indicating subchorionitis. There was no spread of the neutrophils into the chorion or amnion. A mild corresponding foetal inflammatory response was also seen in the umbilical arteries and veins of one of the placentas (CS03). The placenta of CS05 had signs of maternal-foetal malperfusion, and this placenta was also hypoplastic at 389g at 38+0 weeks gestation). However, CS05 had concomitant gestational diabetes mellitus.

**Fig 1.**
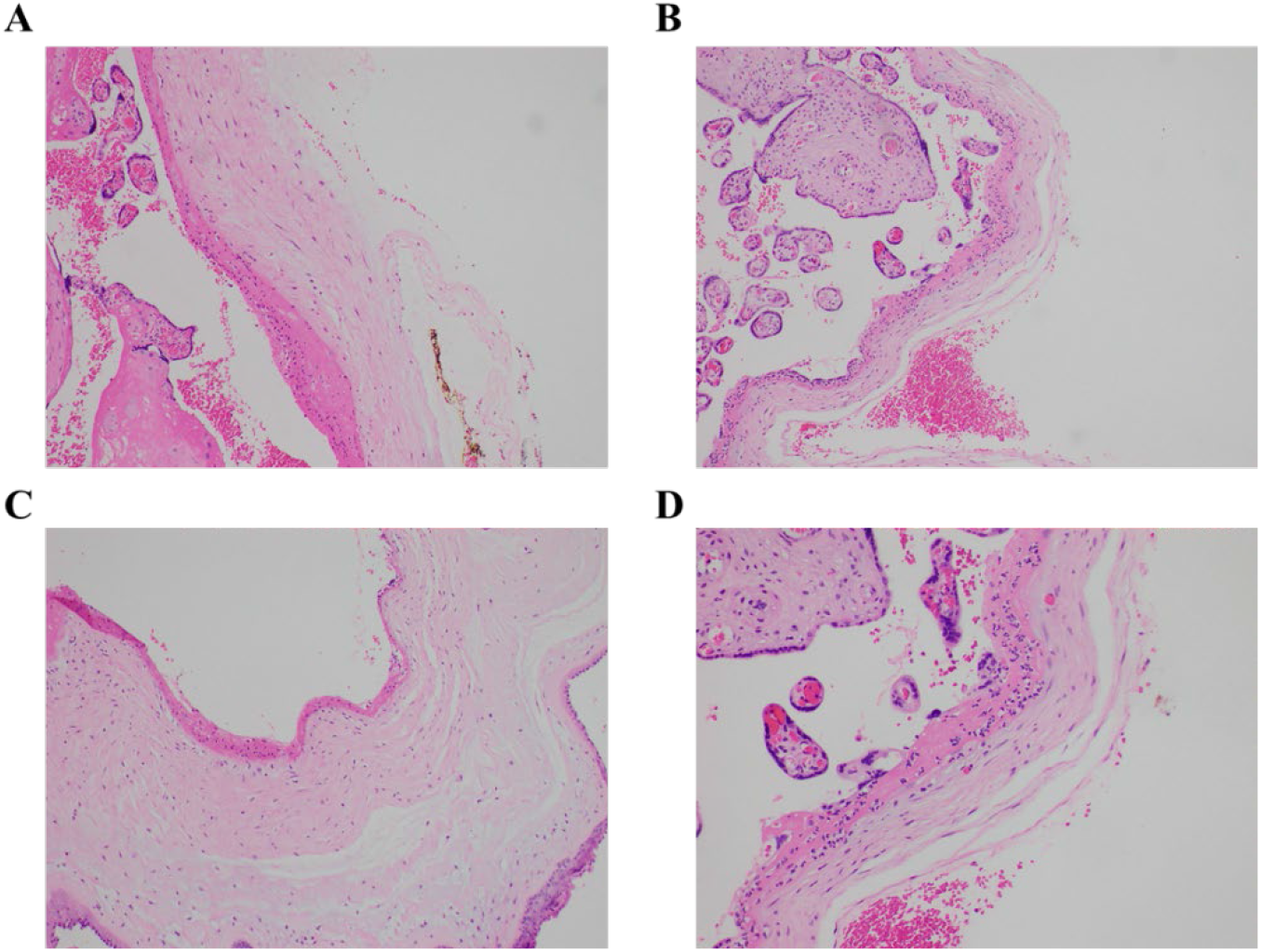
Neutrophilic infiltration present in the placental of convalescent mothers. Microscopic analysis shows a neutrophilic infiltrate in the subchorionic region of the placenta of three patients - (**A**) patient CS2, H&E x 100, (**B**) patient CS3, H&E x100, (**C**) patient CS7, H&E x100. (**D**) Higher power view of the inflammatory process (patient CS2, H&E x200).

### Convalescent mothers carry SARS-CoV-2 specific antibodies in their blood

We first confirmed the presence of IgG antibodies targeting SARS-CoV-2 spike protein (Fig 2A) and spike protein RBD (Fig 2B) in all mothers’ plasma 1-month post-partum. Plasma IgA antibodies targeting the same antigens were expressed more variably; they were detected in ∼50% of the convalescent mothers (Fig 2C, D).

**Fig 2.**
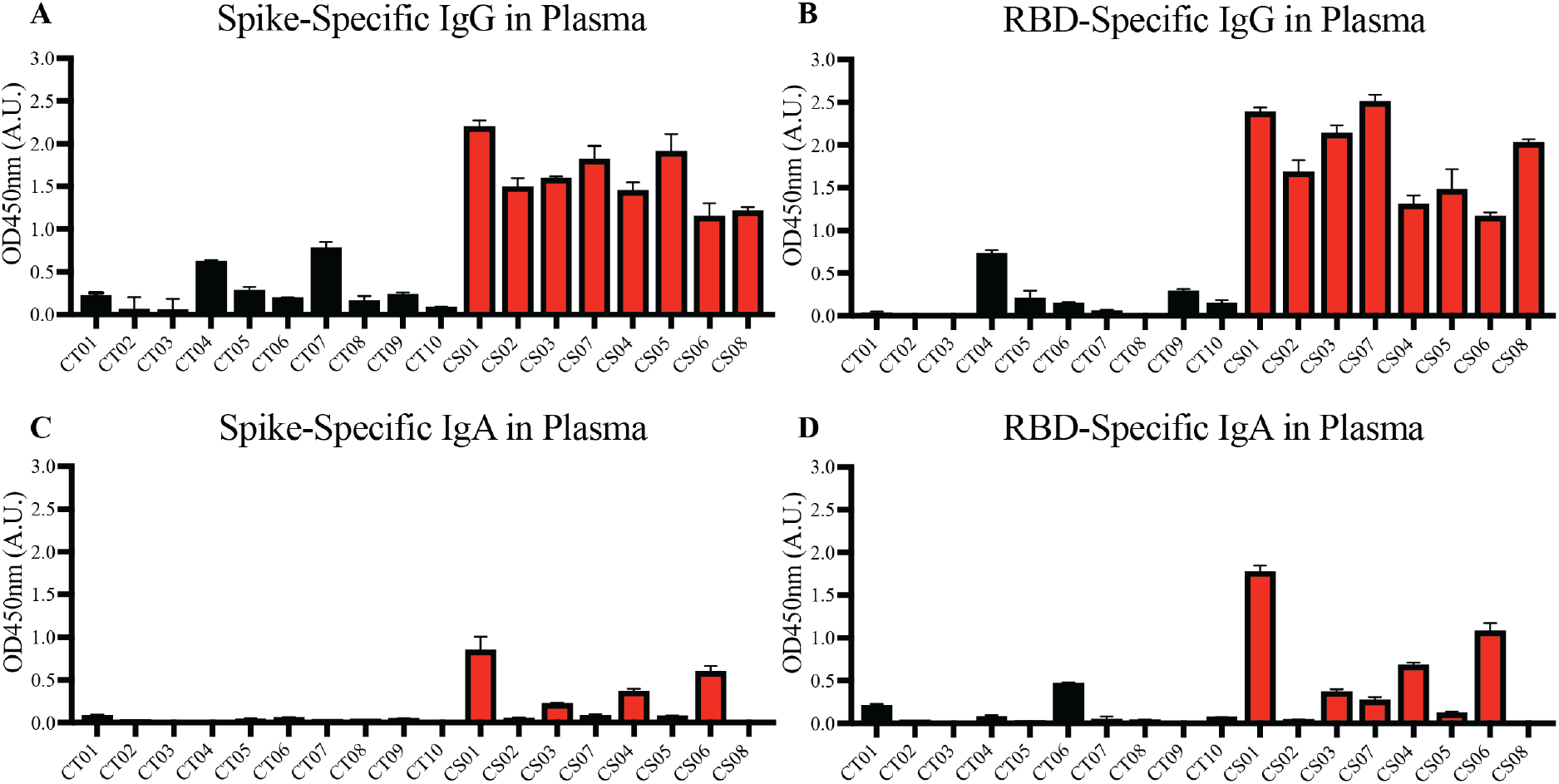
Spike and RBD-specific IgG and IgA present in maternal plasma of convalescent mothers. Using a protein-based ELISA, maternal plasma from control (CT01- CT10) and convalescent (CS01- CS07) mothers obtained 1-month post-partum were used for the detection of spike (**A** and **C**) and RBD (**B** and **D**) specific IgG and IgA antibodies. The bar graphs represent the normalized average signals of antibodies binding to the respective proteins. Convalescent mothers (CS samples) are arranged in ascending order according to their time from COVID diagnosis to delivery.

Most convalescent maternal plasma were able to neutralize SARS-CoV-2 pseudoviruses, albeit to varying extents (Fig 3). Interestingly, plasma from one mother – CS04 – showed no detectable pseudovirus neutralizing ability despite spike- and RBD-specific IgG antibodies being present and comparable to other convalescent mothers. These antibodies are likely to confer some degree of protection against SARS-CoV-2 infections. It is worth noting that neutralization capacity did not correlate with the length of time between disease resolution and point of sampling. In other words, the women who were infected at an earlier stage of pregnancy did not necessarily produce lower titers as one would expect due to antibody decay and contraction of the humoral response.

**Fig 3.**
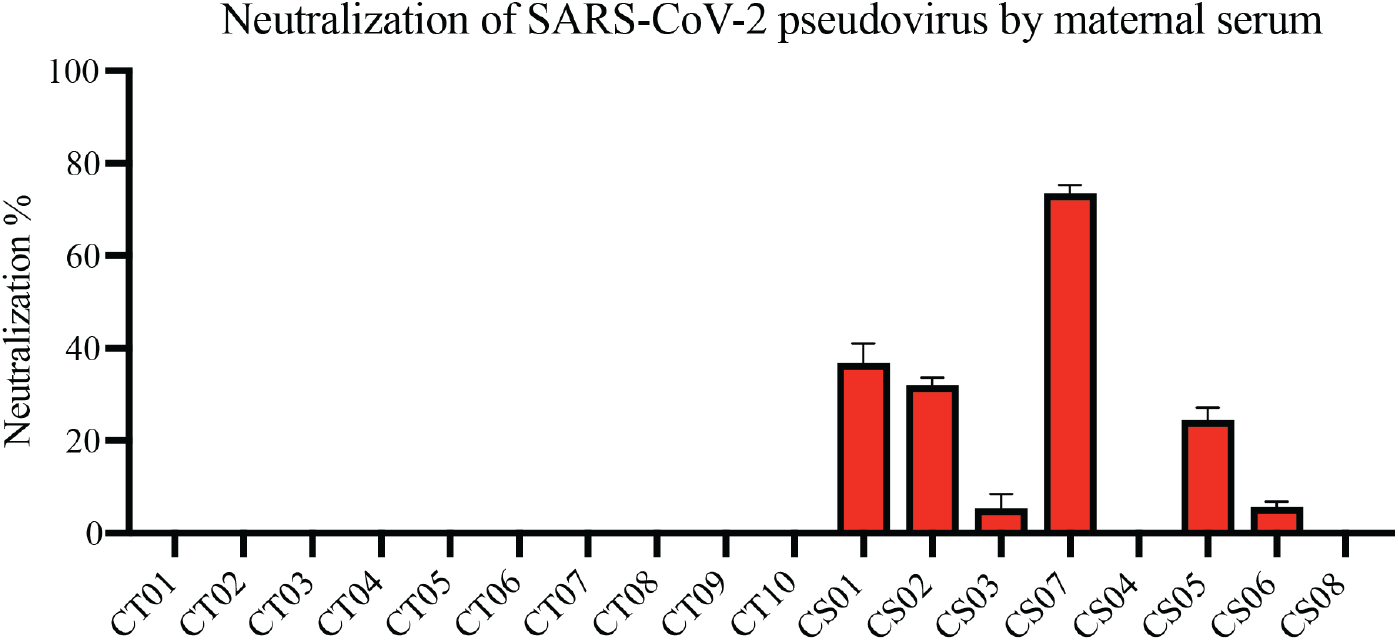
Neutralization of SARS-CoV-2 pseudovirus using maternal plasma. Plasma from control (n=10) and convalescent mothers (n=7). Bar graphs represent the average percentage neutralization. Convalescent mothers (CS samples) are arranged in ascending order according to their time from COVID diagnosis to delivery.

Four immunodominant epitopes S14P5, S20P2, S21P2 and N4P5 were previously identified to be associated with disease severity in adults (age 41 ± 13, years ± standard deviation) [12]. Importantly, antibodies against S14P5 and S21P2 epitopes were able to neutralize SARS-CoV-2-infection [12,13,20]. Hence, we evaluated the presence of antibodies against the four immunodominant epitopes in our cohort. A significant increase in the levels of IgG against S21P2 (Mann Whitney, two tailed, p=0.0205) was observed in the plasma of convalescent mothers compared to controls (S1 Fig C). No significant difference in the levels of IgG against the other three immunodominant epitopes in the plasma of convalescent mothers compared to controls was found (S1 Fig A, B, and D). The IgA levels against the four immunodominant epitopes in convalescent mothers were mostly below the limit of detection. Hence differences in IgA levels in the plasma between both groups cannot be accurately determined (S2 Fig).

### SARS-CoV-2 specific antibodies transferred in-utero to the child wanes over 6 months

An important component of passive immunity is the suite of antibodies transferred across the placenta from mother to child. As shown in Fig 4A and B, compared to the mothers, SARS-CoV-2 specific IgG antibodies were detected in the infants’ sera at comparable or higher levels at birth, showing passive immunity to be successfully transferred via the placenta. We observed a steady decrease in plasma IgG levels in infants from birth to 6 months of age, where the signal approached the lower detection limit. The transplacental transfer ratios for spike- and RBD-specific IgG antibodies ranged from 0.73 to 1.4 6 (Fig 4C and D), in alignment with previous COVID-19 studies and other types of pathogens [21-23].

**Fig 4.**
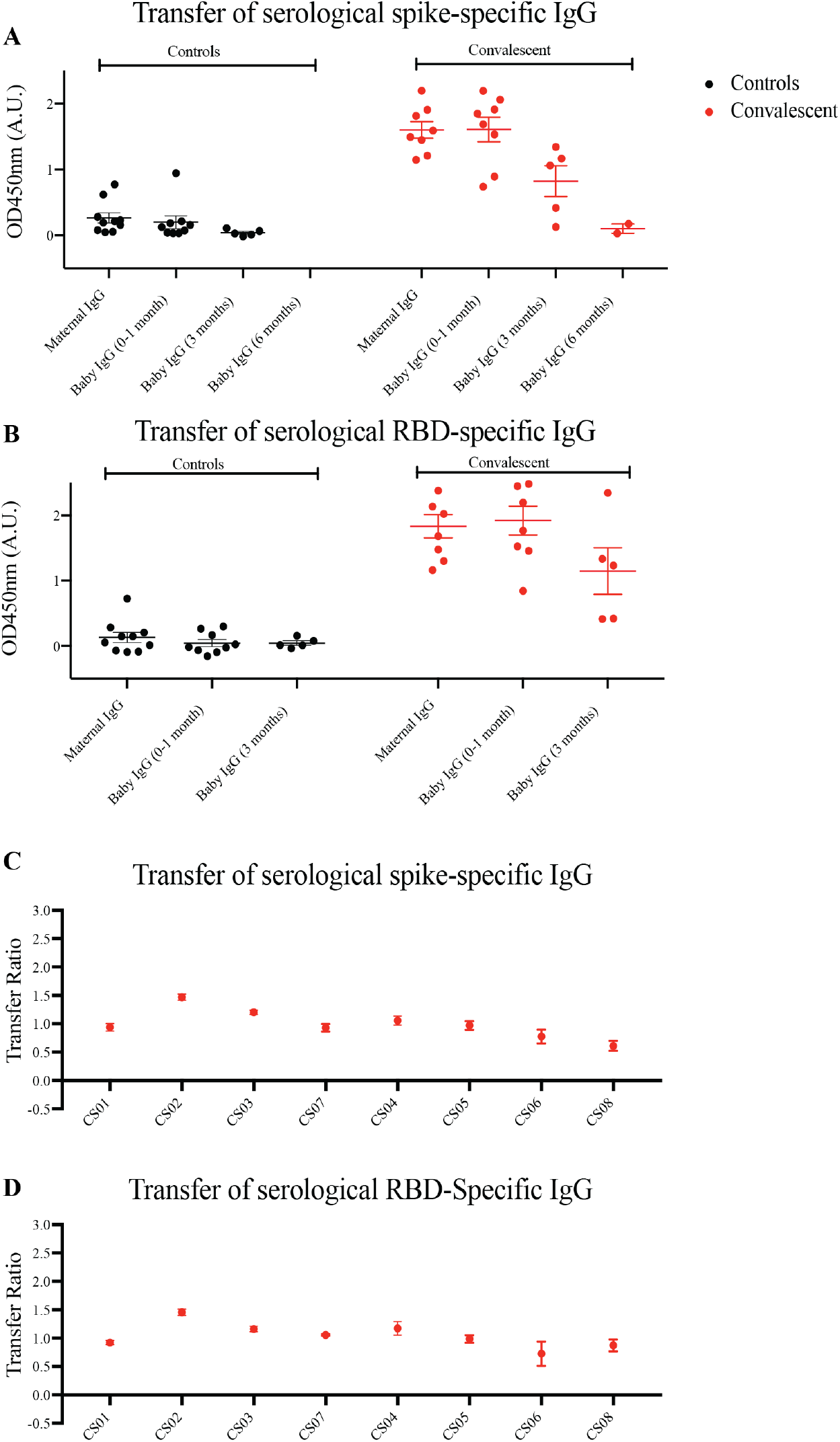
Spike and RBD-specific IgG are passively transferred from convalescent mothers to infants. (**A**) Spike and (**B**) RBD-specific IgG from maternal plasma 1 month post-partum and at three time points from babies’ plasma was determined using ELISA. Transfer ratio at the 1 month timepoint of (**C**) spike and (**D**) RBD-specific IgG from mother to infant was calculated by taking the averaged IgG signal of baby’s over the mother’s IgG. Convalescent mothers (CS samples) are arranged in ascending order according to their time from COVID diagnosis to delivery.

### SARS-CoV-2 specific antibodies are not appreciably secreted into convalescent milk

Apart from transplacental transfer of antibodies, another important mechanism of transmitting passive immunity to the child is via BM. Hence, we sought to measure virus-specific antibody titers in milk of mothers convalescent from antenatal COVID-19. Generally, minimal SARS-CoV-2 spike- and RBD-specific IgG and IgA antibodies were present in milk at 0-3 months postpartum (Fig 5). The same was observed for the epitope-specific antibodies (S3 Fig).

**Fig 5.**
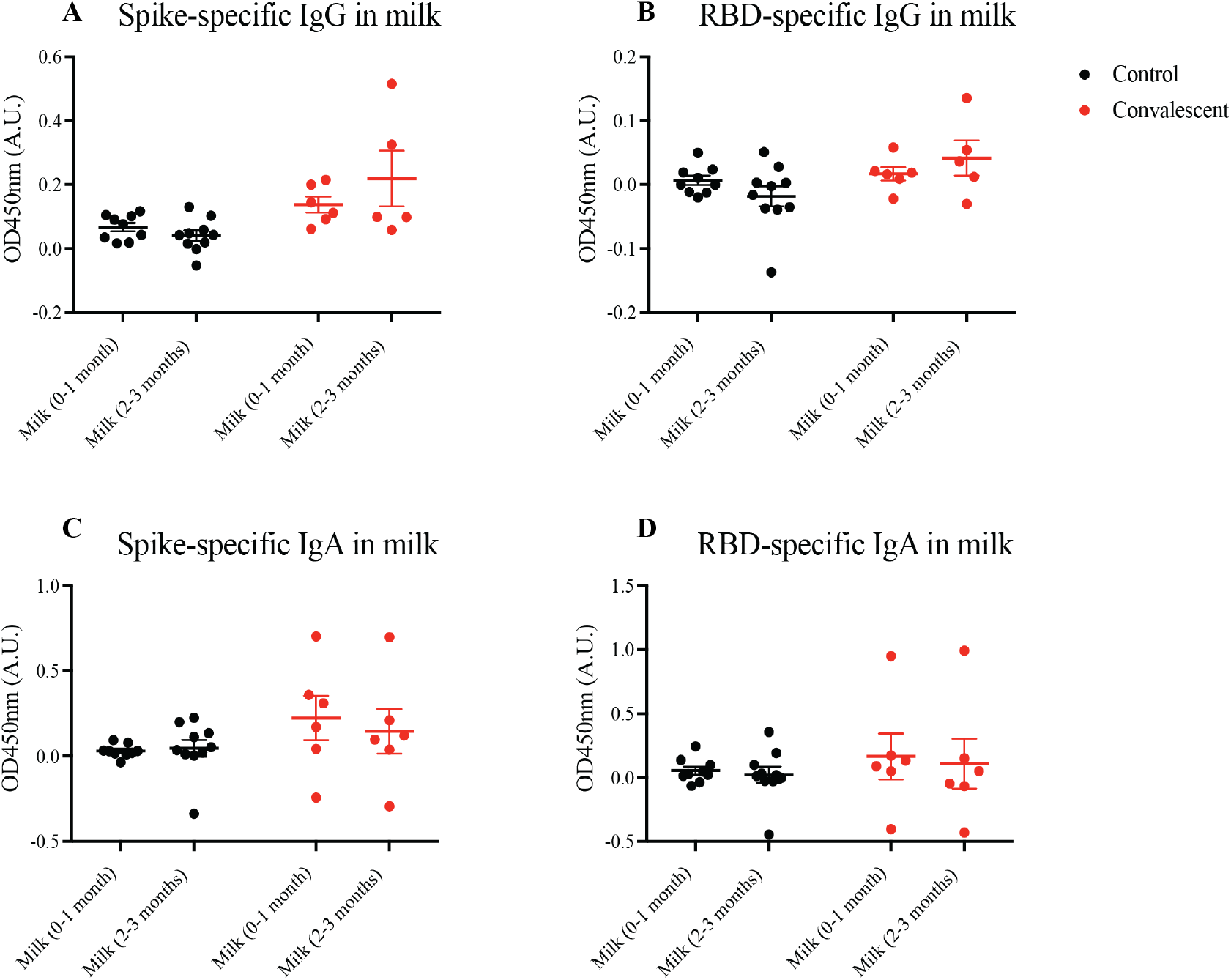
Spike and RBD-specific IgA present in maternal breast milk of convalescent mothers. Using a protein-based ELISA, maternal breast milk from control (CT01- CT10) and convalescent (CS01- CS08) mothers obtained at two time points 0-1months and 2-3months post-partum were used for the detection of spike and RBD-specific IgG and IgA antibodies. Breast milk was screened for (**A**) spike-specific IgG, (**B**) RBD-specific IgG, (**C**) spike-specific IgA, and (**D**) RBD-specific IgA. The bar graphs represent the normalized average signals of antibodies binding to the respective proteins. Convalescent mothers (CS samples) are arranged in ascending order according to their time from COVID diagnosis to delivery.

We noted a singular case, CS04, who produced S21P2-specific IgA responses in plasma and milk that were significantly higher than others, with no decrease observed up to 3 months postpartum (S2 Fig C, S3 Fig C). Notably, CS04 suffers from chronic high-viral load hepatitis C virus (HCV) infection, where the virus is of genotype GT3a. CS04 did not have unusually high serum IgA (Fig 2C and D). Hence, we asked the question whether HCV genotype GT3a encodes S21P2-like antigens that could induce cross-reactive antibody production. Initial Protein BLAST revealed no significant overlap between S21P2 and HCV proteins, including those encoded by GT3a (S1 Appendix). We then compared CS04 against CT10, a control mother who was COVID-19-negative but was HCV genotype GT3a-positive with comparable viral loads. Unlike CS04, CT10 did not have any pre-existing antibodies against S21P2 (S2 Fig C, S3 Fig C). This suggests that the high titers of S21P2-specific IgA in CS04 were not due to cross-reactive IgA-mediated immunity against HCV. It might be that carriage of HCV results in a stronger mucosa-directed anti-viral response marked by higher titers of anti-S21P2 IgA. Nonetheless, we acknowledge the key limitation of small sample size, given the rarity of co-infections by SARS-CoV-2 and HCV in pregnant women; larger studies will be required to clarify interactions or lack thereof between the two viruses.

### Persistence of pro-inflammatory cytokines in convalescent mothers

To gain insight on long-term perturbations in the maternal immune system post-COVID 19, we quantified levels of cytokines with a 41-plex microbead-based immunoassay. Using plasma samples of six control and seven convalescent mothers at 1-month post-partum, we could detect 33 out of 41 cytokines (Fig 6). As far as the assayed cytokines were concerned, both healthy and convalescent mothers had largely similar signatures. Most of the signals for the detected cytokines were not distributed normally, likely due to the fact that they clustered near the lower limit of detection. Notwithstanding, five cytokines, namely interleukin-17RA (IL-17RA), eotaxin, interferon-gamma produced protein-10 (IP-10), monocyte chemoattractant protein-1 (MCP-1) and macrophage inflammatory protein-1β (MIP-1β) exhibited Gaussian distributions and were likely more robust biomarkers. Hence, we focused on these cytokines for further analysis. In order to detect cytokines with increased expression in convalescent mothers, we utilized a statistical threshold of p<0.1 due to small sample size and low power in this study. We found that pro-inflammatory cytokines IL-17RA (p=0.02) and MCP-1 (p=0.0766) were elevated in convalescent mothers than healthy controls, suggestive of a prolonged inflammatory state long after COVID resolution (Fig 6B and C).

**Fig 6.**
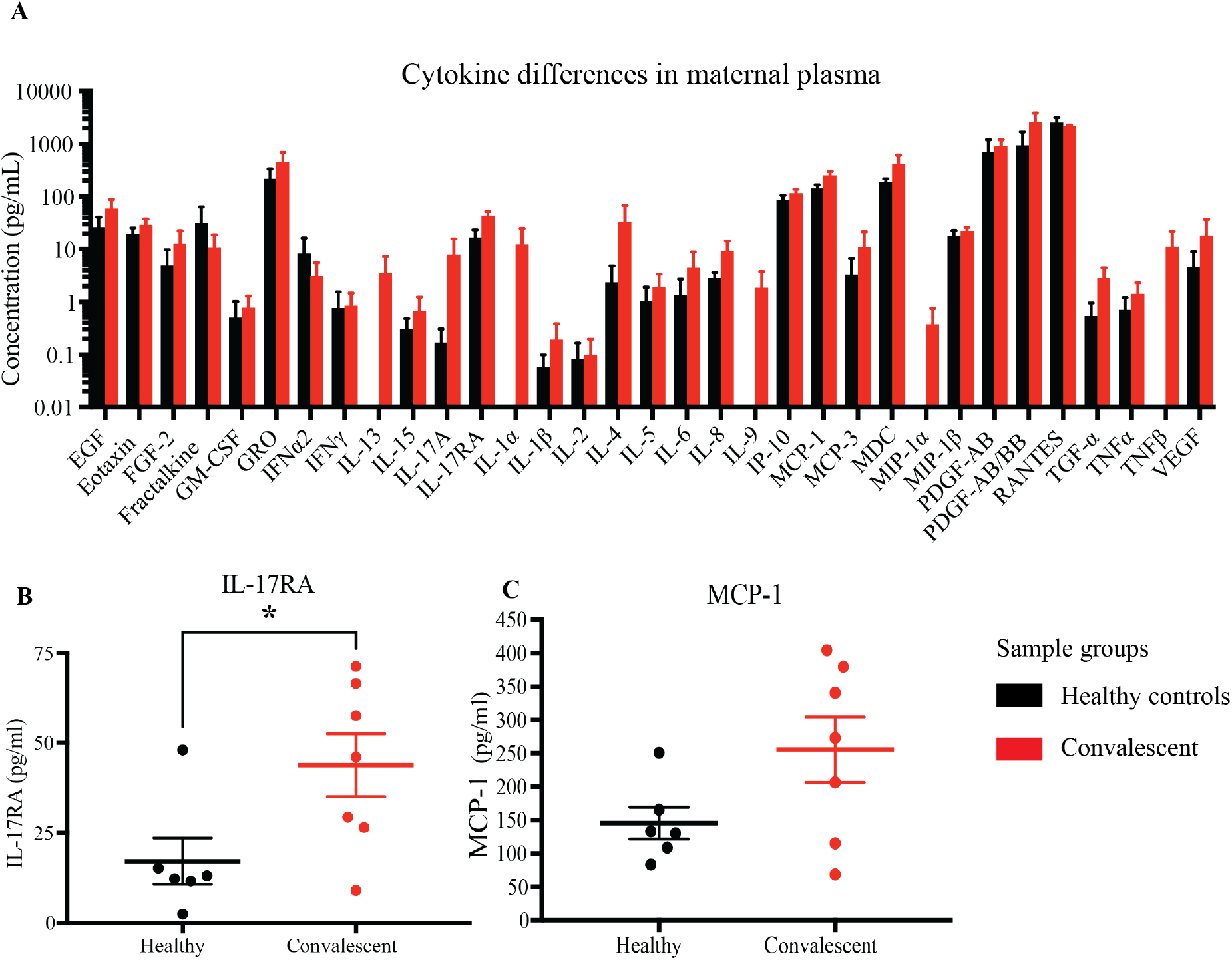
Cytokine profile differences in healthy and convalescent mothers’ plasma. Using 1-month post-partum plasma (n=6 for controls and n=7 for cases), cytokines were evaluated using the 41-plex Luminex™ assay. (**A**) A representation of all cytokines that were detected, (**B**) IL-17RA levels and (**C**) MCP-1. Convalescent mothers are represented in red while healthy controls are represented in black. Bar graphs represent the average log(concentration) from the samples from the respective groups in A while the measured concentrations were used for B and C.

### SARS-CoV-2 specific antibody titers in convalescent milk are boosted with a single dose of Pfizer/BioNTech’s BNT162b2 mRNA vaccine

Given the observation that IgG transferred via the placenta only lasted for about 6 months in the infant, and the scarcity of SARS-CoV-2 specific antibodies in convalescent milk, we investigated alternative methods of passive immunity to protect infants from potential SARS-CoV-2 infection. Three convalescent mothers from our original cohort who were still nursing were given one dose of the BNT162b2 (Pfizer/BioNTech) mRNA vaccine. BM was taken at 4 time points, namely before vaccination, 3 days post-vaccination (PV), 7 days PV and 30 days PV. SARS-CoV-2 specific antibodies were not detectable in milk before vaccination. At 7-30 days PV, both SARS-CoV-2 specific IgG and IgA were generally detected at much higher levels (Fig 7), although CS11 did not secrete detectable levels of RBD-specific IgA into her milk (Fig 7B).

**Fig 7.**
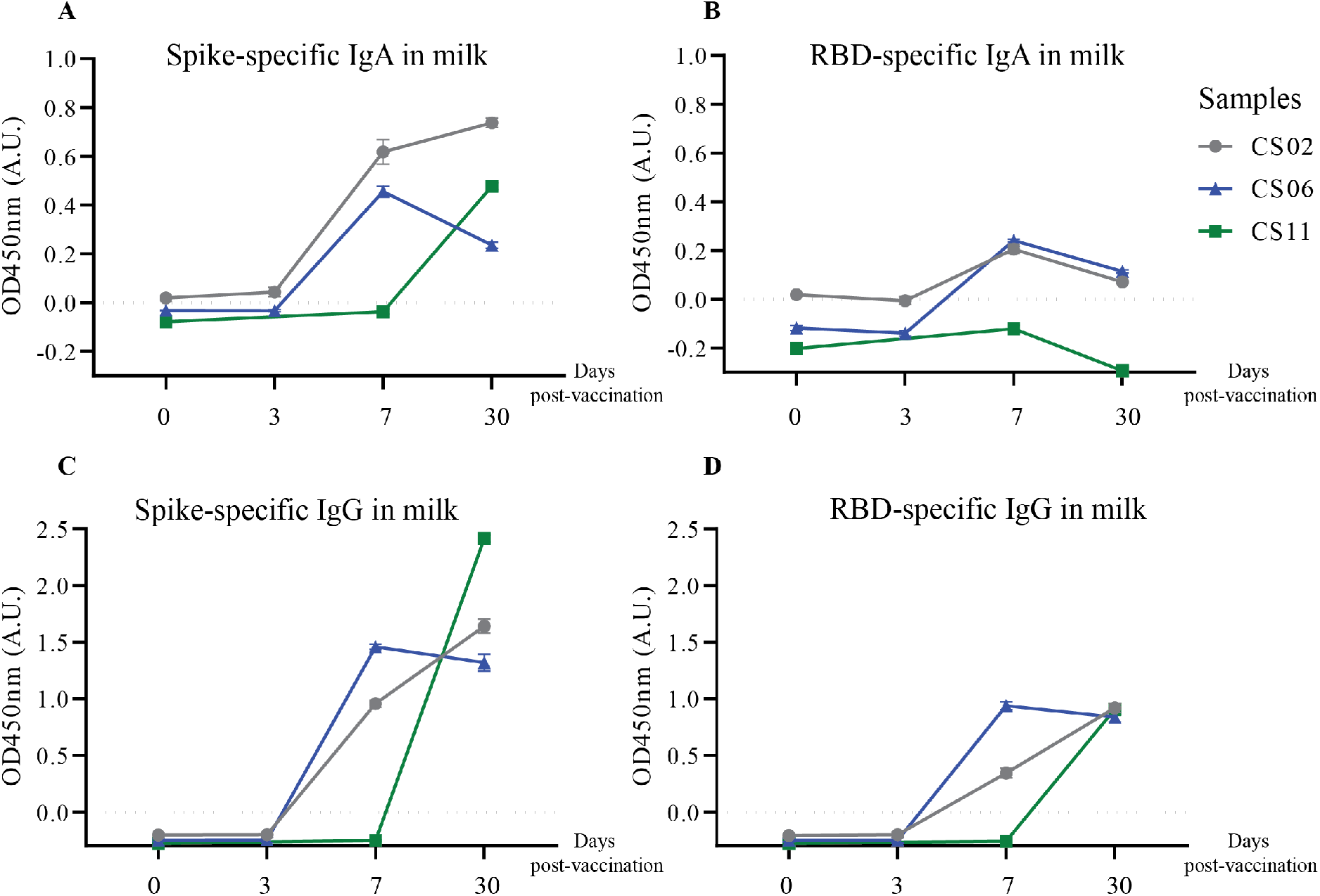
Increased secretion and production of spike-specific IgA and IgG antibodies post-vaccination in convalescent mothers. Breast milk from three nursing convalescent mothers were collected at 4 time points, prior to vaccination (0), 3 days post-vaccination, 7 days post-vaccination, and 30 days post-vaccination for a (**A** and **C**) spike and (**B** and **D**) RBD protein-based IgA and IgG ELISA assay. The x-axis refers to the number of days post-vaccination.

## Discussion

In this study, we show that women who had acquired COVID-19 during pregnancy have high levels of circulating virus-specific IgG in their blood relative to control mothers. Furthermore, spike- and RBD-specific IgG levels in maternal circulation were maintained at relatively high levels regardless of gestation when SARS-CoV-2 infection was acquired in pregnancy; virus-specific IgG could be detected in infants at birth when mothers had acquired antenatal COVID-19 as early as 15 weeks.

However, circulating spike- and RBD-specific IgG levels in infants born to convalescent COVID-19 mothers declined to negligible levels by three months of life. These findings are all consistent with the known patterns of acute gestational infections and passive immunity.

While virus-specific IgG was robustly expressed in convalescent plasma, virus-specific IgA levels in the blood were far more variable. IgA is a marker of acute infection and wanes just as IgG expression starts to pick up during disease resolution [24,25]. Hence, the expectation is that the shorter the interval between virus clearance and the point of sampling, the higher the IgA titers measured at the time of sample collection. We did not see such a clear pattern, which may be due to our small sample size and other factors related to host genetics and the environment that may modulate the plasma IgA response.

It is worth noting that, of the placental samples examined, CS02, CS03 and CS07 showed up abnormal findings indicative of acute inflammation, more than 50 days after their nasopharyngeal swab tested negative by PCR. These findings are broadly consistent with previous studies documenting acute inflammatory pathology, maternal vascular malperfusion, fibrinoid changes and macrophage infiltration [26,27]. Most of the literature on placental pathology in the context of COVID-19 in pregnancy look mainly at acute perinatal infection - trophoblast necrosis and chronic histiocytic intervillositis are common in cases of vertical transmission, whereas materno-foetal malperfusion and villitis are common where there is no vertical transmission. To our knowledge, there are no reports of placental pathology in women convalescent from antenatal COVID-19 at the point of delivery. Interestingly, the four convalescent mothers whose placentae showed abnormal characteristics also showed the lowest titers of virus-specific IgA in their blood. Plasma IgA is predominantly monomeric and capable of engaging FcαRI (also known as CD89) on myeloid cells, presumably to trigger the first line of anti-viral defense. One may speculate that placental granulocytes perform a largely sentinel role to prevent compromise of the placental barrier, and in the absence of high amounts of plasma IgA indicative of active infection, they might be continually and preferentially mobilized to the placenta leading to low levels in the circulation [28].

Inefficient transplacental antibody transfer has been previously reported in a small cohort study of COVID-19 mothers [22]. However, we did not see this particular effect in our cohort, even in the mothers who acquired COVID-19 in the third trimester and whose placentae exhibited signs of inflammation and potential tissue damage. These discrepancies might be due to differences in severity stratification (ratio of asymptomatic: mild: moderate: severe was 6:9:3:4 in Atyeo *et al*. versus 1:6:1:0 in our cohort) and symptom onset to delivery (median of 30.5 days in Atyeo *et al*. versus 98.5 days in our cohort). Also, transplacental transfer occurs most efficiently late into the third trimester, and so infections occurring and resolving earlier in the first and second trimesters are less likely to impact transfer [29].

In our study, we noted a significant elevation of IL-17RA and a trend of increased MCP-1 in convalescent mothers relative to controls (Fig 6B and C). IL-17RA is a soluble receptor of IL-17A, thereby preventing the latter’s association with cell surface receptors. IL-17A has been implicated in SARS-CoV-2 pathogenesis with elevated peripheral IL-17 levels and Th17 infiltration in the lungs of COVID patients [30,31]. The persistence of IL-17A together with other inflammatory cytokines such as IL-12p70, stem cell factor, and IL-1β was discovered in patients up to 180 days post-infection [32]. In agreement with our findings, COVID-19 patients were found to have increased IL-17RA levels during the acute phase, especially in those with milder symptoms [33]. Hence, enhanced IL-17RA may have a protective role in limiting the downstream effects of IL-17; elevated IL-17RA levels may be a prognostic marker of mild disease in pregnant and lactating women. MCP-1 is the other cytokine that was slightly elevated in convalescent mothers relative to control mothers. It is produced by monocytes and macrophages to regulate the migration and infiltration of natural killer cells, monocytes, and T cells [34]. Greater MCP-1 expression characterizes patients with mild COVID-19, suggestive of potential roles in mitigating severity [35]. However, we note that the lack of severe cases in our cohort limits us from investigating whether high IL-17RA and MCP-1 levels are true predictors of mild disease.

In breast milk, low levels of spike-specific IgG were secreted into 2 out of 8 convalescent women’s BM. This is consistent with the existing dogma that IgG is rarely released into milk; the dominant isotype is IgA. Even so, virus-specific IgA was only present in a subset of convalescent mothers’ milk samples. This is somewhat expected as IgA levels are not maintained throughout infection and have been found to be lowest on the outset in mild cases, where most of the subjects in our cohort fall under [24].

We further delineated the humoral response to linear B cell epitopes that have been previously described to be immunodominant. There are certain advantages in investigating these epitopes instead of conformational ones – (1) the system is considerably cheaper to set up for further validation studies, and (2) it is easier to prime cells with peptides which can be synthesized conveniently and do not require complex purification and refolding protocols. We found that most linear epitopes were generally not well-recognized by antibodies present in sera and milk from convalescent mothers. This might be due to the fact that long periods of time had passed between COVID-19 recovery and sampling points; the antibody levels waned, similar to that for other infections. One notable exception is S21P2, where epitope-specific IgG was significantly elevated in convalescent plasma compared to controls. In particular, CS04 produced extremely high levels of S21P2-specific IgA in the BM by mechanisms unknown to us, although preliminary analysis has excluded HCV antigen cross-reactivity as one possible factor. Nonetheless, much of these signals were non-specific and could also be detected in controls who had non-SARS-CoV-2 coronavirus infection or non-coronavirus antenatal URTI.

Given that convalescent sera do contain high levels of spike- and RBD-specific IgG and yet exhibit low, variable neutralization capacities, we infer that most of the IgG are poorly functional in terms of blocking receptor binding, which is consistent with a previous report showing low neutralizing antibody titers in mild cases [36]. The current design of the mRNA vaccines utilizes the 2P mutated spike to stabilize its pre-fusion conformation. As these convalescent women had not received the vaccine prior to infection, their immune systems likely encountered virions that had a mix of pre-fusion and post-fusion trimers, the latter having evolved to perhaps fulfil immunoevasive functions [37]. The consequence for these individuals is the production of weakly neutralizing antibodies that primarily target the post-fusion conformational epitopes. Had the infected individuals produced strongly neutralizing antibodies, those antibodies would have been predominantly raised against the pre-fusion conformation. Given that a single dose of the Pfizer mRNA vaccine was sufficient to boost virus-specific IgG and IgA responses in mothers who had long convalesced from natural SARS-CoV-2 infection, we speculate that the booster-elicited antibodies are directed against both the pre- and post-fusion forms; recall responses are activated in cells recognizing the post-fusion spike, and primary responses are generated in response to the vaccine-encoded pre-fusion spike. Together, these findings underscore the importance of receiving the vaccine even after recovery from natural infection. Additionally, given the lack of safety and efficacy data in administering vaccines to infants, breastfeeding after vaccination could be a viable alternative to conferring some form of mucosal immunity to the vulnerable children, after the loss of transplacentally transferred antibodies at 3 months of age.

A key limitation of our study is the cohort size. Most of our samples were collected early on in the pandemic before Singapore introduced a slew of effective public health measures to stymie the spread of the virus, including travel restrictions, social distancing, and a lockdown. Due to this unique situation, relatively few people were infected in the first wave from April to August 2020. Limited community transmission between August 2020 and August 2021 prevented further subject recruitment and sample collection.

In the context of the present situation, we note two caveats, namely the current dominance of Delta and Omicron over early pandemic variants and the high rate of vaccine uptake by the resident population (∼92% as of 31^st^ March 2022). These two factors preclude meaningful head-to-head comparisons of more recent studies against our cohort in the GIFT study where immunologically naïve individuals were infected with variants possessing lower immunoevasive abilities.

Notwithstanding these differences, our studies provide insights into the pathogenesis of COVID-19 in the under-studied demographics of pregnant and lactating women as well as infants born to them.

## Data Availability

All data produced in the present study are available upon reasonable request to the authors.

## Author contributions

Y.G. – Formal analysis, Investigation, Methodology, Validation, Writing – Original draft preparation, Writing – review & editing. J.M.L. – Conceptualization, Formal analysis, Funding acquisition, Investigation, Project administration, Resources, Validation, Writing – Original draft preparation, Writing – review & editing. J.S.Y.T. –Formal analysis, Investigation, Methodology, Visualization, Writing – Original draft preparation, Writing – review & editing. M.S.F.N – Formal analysis, Investigation, Validation, Writing – Original draft preparation, Writing – review & editing. L.F.P.N – Methodology, Writing – review & editing. B.N. – Investigation, Methodology, Validation, Writing – review & editing. R.G. – Investigation, Writing – review & editing. P.A.M – Resources, Supervision, Writing – review & editing. Z.A. – Resources, Supervision, Writing – review & editing. L.Y.L. – Funding acquisition, Supervision, Writing – review & editing. D.W.Q.L – Formal Analysis, Investigation, Methodology, Validation, Supervision, Writing – review & editing. L.P.C.S – Supervision, Writing – review & editing. Y.Z. – Formal analysis, Investigation, Methodology, Project administration, Supervision, Validation, Writing – Original draft preparation, Writing – review & editing. L.W.W. – Formal analysis, Investigation, Methodology, Project administration, Supervision, Validation, Writing – Original draft preparation, Writing – review & editing.

All authors declare that they have no conflicts of interest.

## Acknowledgements

This work was funded by National University Hospital of Singapore (NUHS) seed grants awarded to J.M.L. and L.Y.L. L.W.W. is a recipient of the Singapore National Medical Research Council Open Fund Young Investigator Grant (NMRC OF-YIRG; Grant number MOH-000545-00) and an award from the A*STAR Biomedical Research Council (BMRC) Central Research Fund for Use-Inspired Basic Research (CRF-UIBR).

We thank Antibody Engineering Programme, Life Sciences Institute, NUS for providing the purified spike protein and RBD. We thank Associate professor Tan Yee Joo from Department of Microbiology and Immunology, Yong Loo Lin School of Medicine, NUS for the ACE2 stably expressing CHO cells and plasmid encoding SARS-CoV-2 S protein for the pseudotyped lentiviral production. We thank the participating mothers and children for their kind donation of samples.

## Notes

### Competing Interest Statement

The authors have declared no competing interest.

### Funding Statement

This study was funded by National University Hospital of Singapore (NUHS) seed grants awarded to J.M.L. and L.Y.L. L.W.W. is a recipient of the Singapore National Medical Research Council Open Fund Young Investigator Grant (NMRC OF-YIRG; Grant number MOH-000545-00) and an award from the A*STAR Biomedical Research Council (BMRC) Central Research Fund for Use-Inspired Basic Research (CRF-UIBR).

### Author Declarations

IRB of National Healthcare Group gave ethical approval for this work (Gestational Immunity For Transfer GIFT: DSRB Reference Number: 2020/00483). Written informed consent was obtained from all subjects (and where applicable, parents), and the study was conducted in accordance with the Helsinki Declaration.

