## Supplementary Figures and Tables for "Immune and pathophysiologic profiling of antenatal COVID-19 in the GIFT cohort: A Singaporean case-control study"

### Supporting information

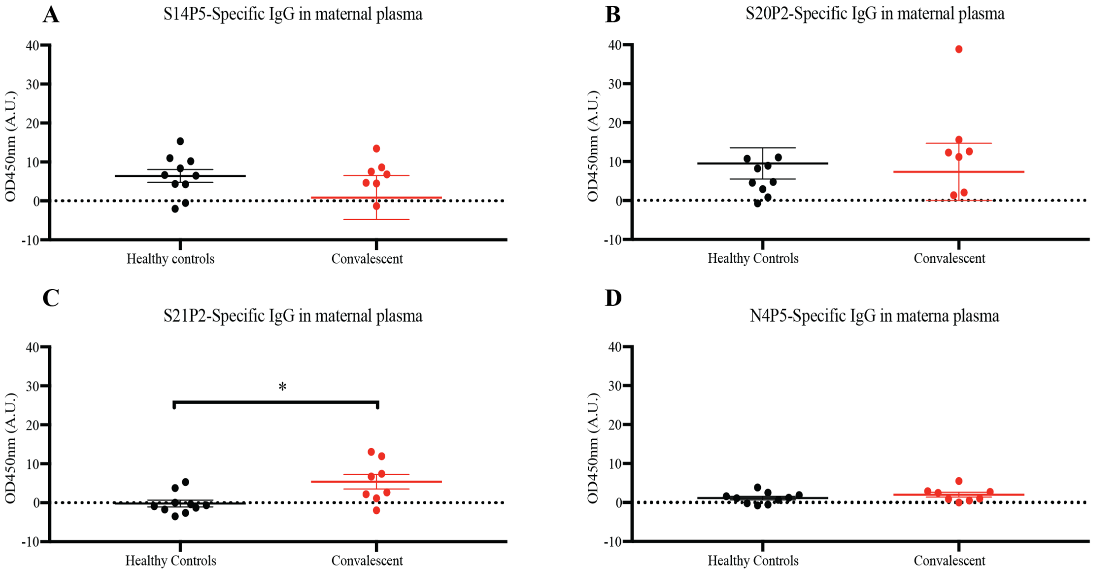


**S1 Fig. Screening IgG antibodies against four immunodominant epitopes in maternal plasma**. Using 1-month post-partum maternal plasma from control (CT01-CT10) and convalescent (CS01-CS08) mothers. An evaluation of IgG levels against (**A**) S14P5, (**B**) S20P2, (**C**)S21P2, and (**D**) N4P5 peptides. Bar graphs represent the normalized average signals, and the dotted line is the averaged signals from the controls.

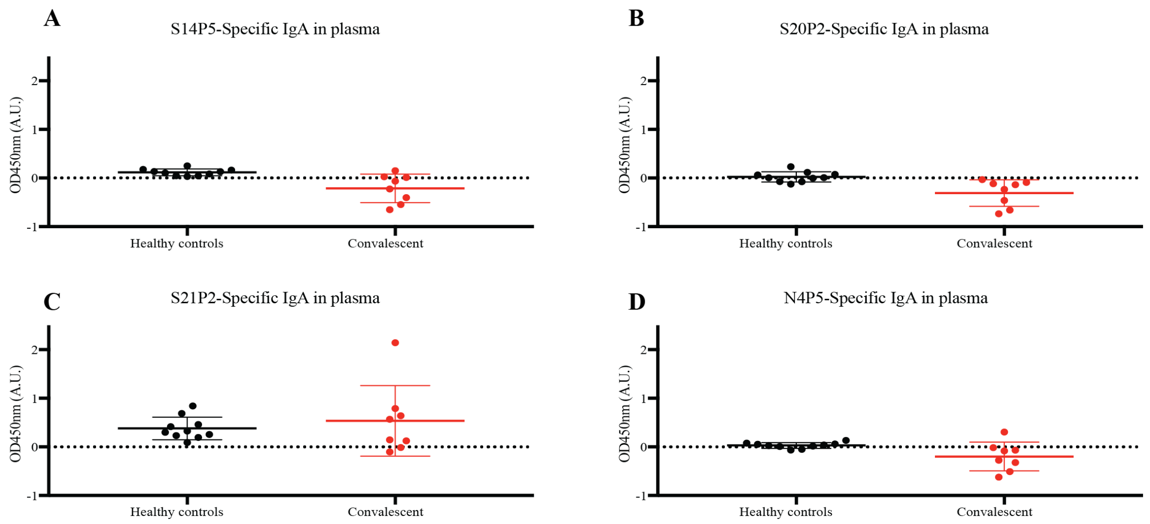


**S2 Fig. Screening IgA antibodies against four immunodominant epitopes in maternal plasma**. Using 1-month post-partum maternal plasma from control (CT01-CT10) and convalescent (CS01-CS08) mothers. An evaluation of IgA levels against (**A**) S14P5, (**B**) S20P2, (**C**)S21P2, and (**D**) N4P5 peptides. Bar graphs represent the averaged normalized signals, and the dotted line is the averaged signals from the controls. The sample with the highest signal in (C) under convalescent mothers represents CS04.

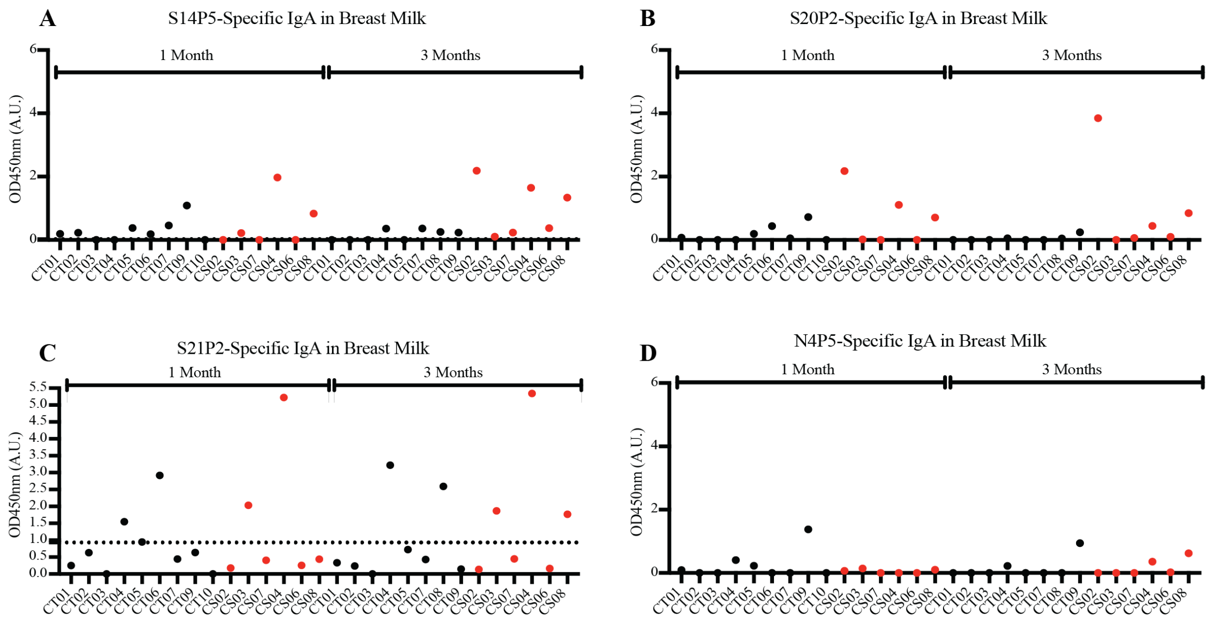


**S3 Fig. Screening IgA antibodies against four immunodominant epitopes in maternal breast milk at 1 and 3 months post-partum.** Using 1 and 3-month post-partum maternal breast milk from control (CT01-CT10) and convalescent (CS01-CS08) mothers. An evaluation of IgA levels against (**A**) S14P5, (**B**) S20P2, (**C**) S21P2, and (**D**) N4P5 peptides. Bar graphs represent the averaged normalized signals, and the dotted line is the averaged signals from the controls. Convalescent mothers (CS samples) are arranged in ascending order according to their time from COVID diagnosis to delivery.

**S1 Table. Information on the time from diagnosis to delivery for the convalescent participants.**

| Sample | Interval from COVID diagnosis to delivery (Days) |
|---|---:|
| CS01 | 37 |
| CS02 | 69 |
| CS03 | 76 |
| CS04 | 105 |
| CS05 | 111 |
| CS06 | 226 |
| CS07 | 92 |
| CS08 | 250 |

**S2 Table. Individual characteristics of convalescent women.**

| | CS01 | CS02 | CS03 | CS04 | CS05 | CS06 | CS07 | CS08 |
|---|---|---|---|---|---|---|---|---|
| Maternal Age Range (Years) | 26-30 | 26-30 | 36-40 | 31-35 | 26-30 | 31-35 | 26-30 | 26-30 |
| Maternal Ethnicity | Indian | Caucasian | Caucasian | Malay | Malay | Chinese | Indian | Malay |
| Diagnosis of COVID-19 (Gestation, weeks) | 36 | 32 | 29 | 24 | 22 | 10 | 28 | 4 |
| COVID severity (WHO classification) | Mild | Mild | Mild | Mild | Mild | Moderate | Asymptomatic | Mild |
| Mode of delivery | Vaginal | Vaginal | Vaginal | Vaginal | Vaginal | Vaginal | Vaginal | Vaginal |

**S3 Table. Individual laboratory results of convalescent women peri-delivery**

| | CS01 | CS02 | CS03 | CS04 | CS05 | CS06 | CS07 | CS08 |
|---|---|---|---|---|---|---|---|---|
| COVID-19 nasopharyngeal swab (closest swab done prior to delivery) | Negative | Negative | Negative | Negative | Negative | Not done | Not done | Not done |
| COVID-19 in serum | Negative | Negative | Negative | Negative | Negative | Negative | Negative | Negative |
| COVID-19 in vaginal swab | Negative | Negative | Not done | Negative | Not done | Negative | Negative | Negative |
| COVID-19 swab of umbilical cord | Negative | Negative | Negative | Negative | Negative | Negative | Negative | Negative |
| COVID-19 swab of placenta | Negative | Negative | Negative | Negative | Negative | Negative | Negative | Negative |
| COVID-19 swab of amniotic fluid | Not done | Negative | Not done | Negative | Negative | Negative | Negative | Not done |

| | | | | | | | | |
|---|---|---|---|---|---|---|---|---|
| COVID-19 swab of breast milk | Not done | Negative | Negative | Negative | Not done | Negative | Inconclusive | Not done |

**S1 Appendix. Protein BLAST result between S21P2 peptide and HCV proteins**
